# Characterising the neurobiological mechanisms of action of exercise and cognitive behavioural interventions for rheumatoid arthritis fatigue: an MRI brain study

**DOI:** 10.1101/2023.08.23.23294366

**Authors:** Amir Dehsarvi, Salim Al-Wasity, Kristian Stefanov, Stewart Wiseman, Stuart Ralston, Joanna Wardlaw, Richard Emsley, Eva-Maria Bachmair, Jonathan Cavanagh, Gordon D. Waiter, Neil Basu

## Abstract

**Background:** Chronic Fatigue is a major clinical unmet need among patients with Rheumatoid Arthritis (RA). Current therapies are limited to non-pharmacological interventions, such as personalised exercise programmes (PEP) and cognitive behavioural approaches (CBA), however, still most patients continue to report severe fatigue. To inform more effective therapies, we conducted an MRI brain study of PEP and CBA, nested within a randomised controlled trial (RCT), to identify their neurobiological mechanisms of fatigue reduction in RA.

**Methods:** A sub-group of RA subjects (n=90), participating in a RCT of PEP/CBA for fatigue, undertook a multi-modal MRI brain scan following randomisation to either usual care (UC) alone or in addition to PEP/CBA, and again after the intervention (6 months). Brain regional volumetric, functional, and structural connectivity indices were curated and then computed employing a causal analysis framework. The primary outcome was fatigue improvement (Chalder Fatigue Scale).

**Findings:** Several structural and functional connections were identified as mediators of fatigue improvement in both PEP and CBA compared to UC. PEP had a more pronounced effect on functional connectivity than CBA, however, structural connectivity between the left isthmus cingulate cortex (L-ICC) and left paracentral lobule (L-PCL) was shared and the size of mediation effect ranked highly for both PEP/CBA (ß_Average_=-0·46, SD 0·61; ß_Average_=-0·32, SD 0·47, respectively).

**Interpretation:** The structural connection between the L-ICC and L-PCL appears to be a dominant mechanism for how both PEP/CBA reduces fatigue among RA patients. This supports its potential as a substrate of fatigue neurobiology and a putative candidate for future targeting.

## Introduction

Fatigue is pervasive among people with inflammatory rheumatic diseases ^1^. In rheumatoid arthritis (RA), for example, 80% of patients report significant fatigue ^2^ and over 70% consider this equal to pain in terms of burden ^3^. Critically, most patients continue to experience severe fatigue despite successful anti-inflammatory treatment of their underlying disease ^4^. This common scenario represents one of the principal challenges to face rheumatologists in routine practice. Management is currently limited to exercise and psychosocial interventions ^5^. Although programmes of these non-pharmacological therapies have been successfully implemented into rheumatology services ^6^, their clinical effects are generally small to medium in size, with most recipients still reporting significant levels of fatigue. By understanding the mechanisms of fatigue reduction of these treatments, more effective interventions can be developed in the future.

Epidemiological investigations implicate the importance of brain factors (e.g., mental health) rather than with peripheral measures (e.g., inflammation)^7^ as a focus for putative fatigue mechanisms. Delineating in vivo human brain mechanisms is restricted by access to the brain, however, imaging offers a non-invasive surrogate approach. In RA, and other chronic diseases, magnetic resonance imaging (MRI) modalities have identified multiple brain correlates of fatigue. These characterise the brain beyond what is achievable with conventional macroscopic clinical scans. They include volumetric morphometry, which enables quantification of regional volumes, cortical thickness, and surface areas; diffusion tensor imaging (DTI), which delineates white matter tracts and subsequent structural connectivity between different brain regions and functional connectivity (fcMRI), an adaptation of functional MRI data that examines intrinsic connectivity. Together, these networks or ‘connectomes’ help map the communications between different brain regions. This is especially relevant in the context of complex behaviours, such as fatigue, whose mechanisms are not likely constrained to a single region.

These MRI modalities have consistently associated fatigue to frontal, parietal, and cingulate cortices, alongside sub-cortical striatal structures ^8^. In RA, higher fatigue levels were related to stronger functional connectivity between the dorsal attention network (DAN) and bilateral prefrontal cortex as well as greater right putamen volumes ^9^. Notably, MRI brain studies examining the neural effects of exercise and psychosocial interventions, e.g., cognitive behavioural approaches (CBA), have implicated similar regions ^10,11^. Taken together, the neurobiological effects of exercise and psychosocial interventions plausibly modulate fatigue specific brain networks that represent the final common pathway of this heterogeneous symptom. Translational research can focus on probing major network hubs using non-invasive neuromodulation technologies as a basis for novel therapies. However, due to the diffuse nature of established neural correlates, it is uncertain which regions should be the focus for treatment. Moreover, the apparent variability, and often lack of reproducibility, of previously reported brain regions of interest may be attributable to sub-optimal study designs.

Limitations of previous MRI brain fatigue studies include their cross-sectional or uncontrolled longitudinal design, preventing causal inferences. Clinical study designs applying mediation analysis via randomised controlled experiments are considered gold standard in addressing this limitation ^12^. Mediation analyses can examine why observed relations between variables exist or help understand outcomes associated with interventions to illuminate causal mechanism(s) through which variables relate ^13^. The high dimensionality of neuroimaging data has previously precluded mediation analysis, however, advanced computational approaches have now enabled researchers to investigate the mechanism of action of exercise and CBA interventions with view to deriving mechanistic insights into cognitive impairment ^14^. Given their established benefit for fatigue, such interventions could be similarly leveraged, for the first time, to aid the selection and prioritisation of putative neurobiological mediators of this patient priority.

This study is the first to embed multimodal MRI brain scans in a randomised controlled trial (RCT) of exercise and psychosocial therapies for RA fatigue. The parent trial evidenced statistically and clinically important fatigue improvements of both a telephone delivered exercise and CBA intervention compared to usual care. This MRI sub-study aimed to employ mediation analyses to characterise the neurobiological mechanisms of action of these interventions, and then rank the identified putative causal neural regions for future therapeutic targeting.

## Methods

### Study design

The study is a nested 3 Tesla MRI brain sub-study within the Lessening the Impact of Fatigue Trial (LIFT) ^15^.

### Parent Trial

LIFT was a RCT to test the hypothesis that usual care with either telephone delivered CBA, or personalised exercise programme (PEP) is more effective than usual care (UC; a patient education booklet) alone. CBA involved a structured psychological intervention, aiming to replace unhelpful beliefs/behaviours with adaptive ones. Alternatively, PEP targeted intolerance of physical activity and reversal of deconditioning. In total n=368 inflammatory rheumatic disease patients were randomised (RA=202). Participants randomised to an active arm received up to 8 sessions lasting maximum 1hr of therapy over a period of 6 months. The primary outcome was self-reported fatigue at 12 months (measured by Chalder Fatigue Scale) ^6^ – see ^15^ for further details of their characteristics and outcomes.

### MRI Sub-study

**Inclusion Criteria** – subjects must have been a) ≥18 years, b) consented to the parent trial randomisation, c) classified with RA according to the 2010 American College of Rheumatology/European League Against Rheumatism criteria ^16^, d) fatigued > 3 months, e) significantly fatigued (≥6 on 1-10 Visual Analogue Scale), f) considered to have stable RA (as defined by unchanged immunomodulatory therapy in the previous 3 months).

**Exclusion Criteria** – subjects were not considered if they had a) alternative medical explanations for their fatigue, e.g., anaemia, b) contra-indications to MRI, c) already started an intervention.

Recruitment processes have been previously reported ^6^. Randomisation was undertaken using a computer-generated sequence, participants were allocated to receive either of the two treatments or usual care (1:1:1 ratio). Those eligible were provided information on the sub-study and following randomisation were offered an appointment to attend an MRI research facility within a month (and prior to their first telephone consultation if they receive active therapy).

**Clinical Assessment** – All subjects were comprehensively characterised at baseline and 6 months as part of the parent trial. This included disease activity, CRP, disease duration and co-morbidities (Charlson index)^1^.

### MRI Imaging Parameters

MRI multimodal data were acquired in three scanning sites with two system types; a 3 Tesla Philips Achieva X-series (Philips, Best, Netherlands) and a 3 Tesla Siemens PRISMA (Siemens, Erlangen, Germany) using 32 channel phased-array head coils. All patients were consented prior scanning. The multimodal scanning consisted of:

1. **Structural MRI**: Structural MRI data was acquired by a T1-weighted MPRAGE/fast-field echo 3D structural scan with the following parameters: repetition time (TR)=8·2ms, echo time (TE)=3·8ms, inversion time (TI)=1025·7ms, flip angle (FA)=8°, field of view (FOV)=240×240mm, matrix size=240×240 with 160 sagittal slices, voxel size=1×1×1mm^3^, total scan time=5·63min.
2. **Resting-State Functional MRI**: Resting-state functional MRI data was collected with a T2*-weighted gradient-echo echo-planar imaging (EPI) pulse sequence with the parameters: TR=1·95s, TE=26ms, FA=70°, FOV=240×240mm, matrix size=128×128 with 30 transverse slices in ascending order, voxel size=1·88×1·88×3·5mm^3^ with slice gap=1·5mm, 308 volumes, total scan time=10·01 min. Subjects were instructed to keep their eyes open during the scan and focus on a displayed fixation cross.
3. **Diffusion MRI**: Diffusion MRI were acquired using a single-shot spin-echo EPI sequence with the following parameters: TR=7010ms, TE=90ms, FA=90°, FOV=220×220mm, matrix size=96×96 with 60 transverse slices, voxel size=2·3×2·3×2·3mm^3^ with no gap, number of excitations=1, gradient directions=64 (b=2000 s/mm^2^) and 8 volumes of unweighted (b=0) images.

### MRI Data Pre-processing

1. **Structural MRI:** All the structural data were pre-processed using FreeSurfer image analysis suite v6·0 (http://surfer.nmr.mgh.harvard.edu/). The pre-processing pipelines includes skull stripping motion correction, intensity normalisation, Talairach registration, skull stripping, subcortical segmentation and labelling, segmentation of WM, tessellation of the GM/WM and GM/CSF boundaries, automated topology correction, and surface deformation, and cortical surface reconstruction ^17^. Each individual’s brain was parcellated into 84 cortical and subcortical ROIs (42 ROIs per hemisphere). Surface area, cortical thickness and volume measures were extracted from each individual’s ROI.
2. **Resting-State Functional MRI:** All the resting-state fMRI data were pre-processed using CONN (http://www.nitrc.org/projects/conn), a MATLAB-based cross-platform software for pre-processing and analysing MRI functional connectivity. The default surface-based subject-space analyses pipeline was utilised. Briefly, the analysis pipeline consists of motion correction, slice-time correction, outlier detection (scrubbing) using ART toolbox, coregistration to the structural volume, smoothing (8mm FWHM kernel), denoising, nuisance regression (Principal Components of WM and SCF, mean GM signal, six rigid-body realignment movement covariates and scrubbing series), linear detrending and band-pass filtering (0·008-0·09 Hz). First level ROI-ROI connectivity was computed using 84 cortical and subcortical ROIs, and an 84×84 symmetrical Fisher Z-transformed matrix (FC) was estimated using ROIs BOLD signals of each subject.
3. **Diffusion MRI:** All the diffusion data were pre-processed using FMRIB Software package (FSL v6·0 and FSL Diffusion Toolbox (FDT) - https://fsl.fmrib.ox.ac.uk/fsl/fslwiki). The pre-processing procedures includes, skull stripping, eddy current distortion correction, motion correction, Fractional Anisotropy (FA) calculation, probabilistic distributions estimation using The GPU version of BEDPOSTX tool ^18^, and finally performing the probabilistic tractography to estimate the structural connectivity probability among 84 cortical and subcortical regions using the PROBTRACKS tool ^19^, which yields an 84×84 asymmetrical structural connectivity (SC) matrix for each subject.

After pre-processing, all the MRI features from the three treatment subgroups were merged and the difference (Δ) between the two sessions were calculated and a multimodal matrix (ΔMM) was created by horizontally concatenating the above ΔMRI matrices forming a matrix of (N_subj_ × 10,678_features_).

### The Mediation Analysis

An agnostic multi-group multi-mediator mediation analysis was implemented to explore how brain imaging features mediate the relationship between fatigue improvement and each of the two intervention groups (relative to control).

We examined the neural variables of fatigue, which are not necessarily causing fatigue. Nevertheless, inferences for the two concepts are statistically identical ^20,21^. The mediators are neural/brain imaging features that can be used to describe the relationship between the interventions (the independent/exposure variable) and fatigue improvement (the dependent variable/outcome), see Figure 1. The analysis investigates the indirect effect carried by individual mediators separately and was implemented using the *mmabig* package in statistics software R ^22^.

**Figure 1:**
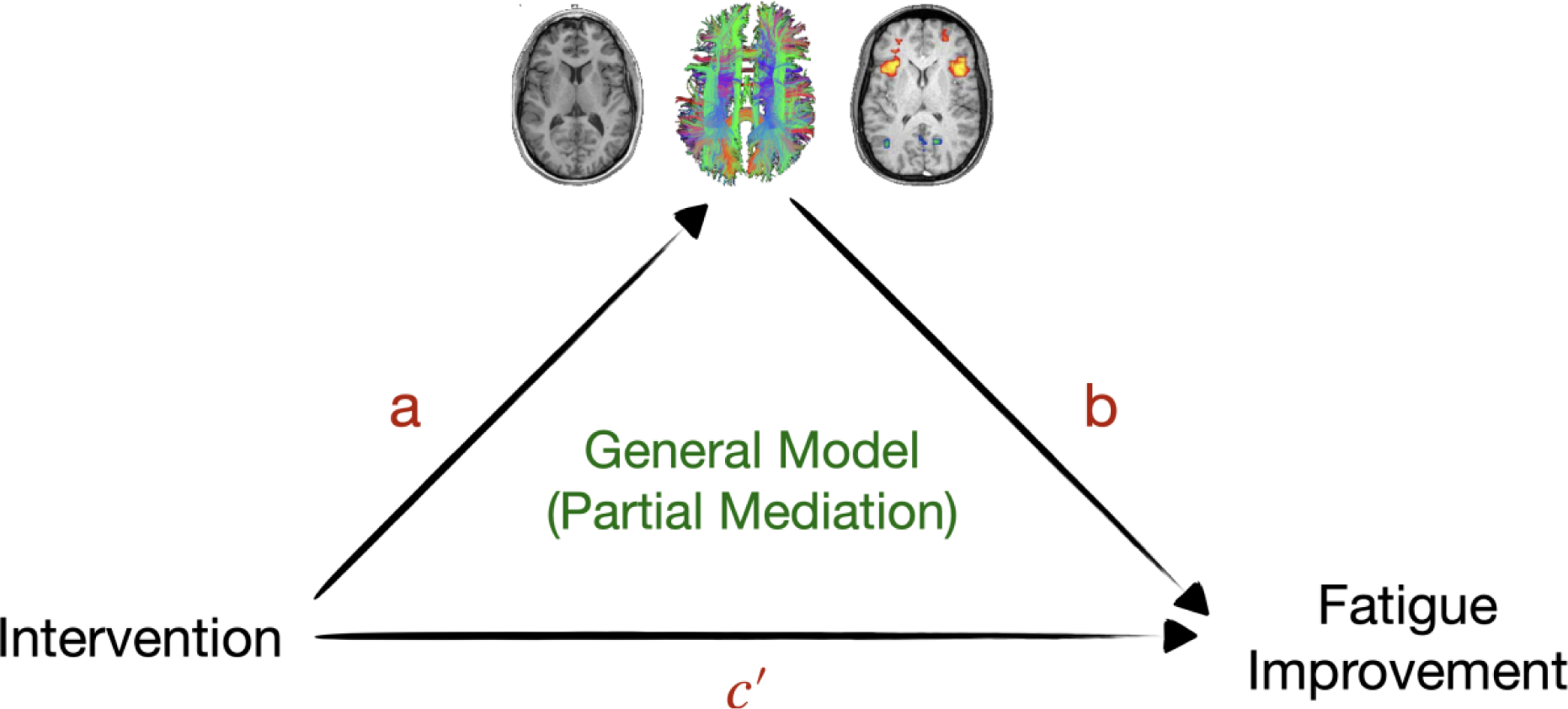
The general mediation model for this study. The brain imaging measures are tested for mediation in fatigue improvement in different intervention groups. The indirect effect is the product of the a and b path coefficients, which measures the changes in the dependent variable when the independent variable is fixed and the mediator variable changes (by the amount that it would have changed if the independent variable increased by one unit). The direct effect is denoted as c^’^ and measures the changes in the independent variable when the dependent variable increases by one unit (the mediator variable remains unchanged) ^23,24^.

The outcome (Y) is Chalder fatigue score improvement over time. The predictor (exposure variable, X) is the intervention group, a (multi-)categorical variable of the values of 1 or 2 or 3. Group 2 (reference group) is the UC receivers. The analysis was blind to the two intervention groups (PEP and CBA). The potential mediators (M) are the neural/brain imaging features. We calculated the difference between the values of the features for session 1 subtracted from the corresponding values for session 2. Furthermore, five different variables as covariates (exogenous variables, denoted as Z) were included in the analysis. The covariates consisted of Chalder fatigue score for session 1, age at session 1, total brain volume for session 1, study centre, and gender. A generalised linear model was used for modelling the relationship among the variables that were included in the mediation analysis and the response. The linkage function was set to gaussian (link = “identity”) ^22^. The regression technique that was used in this research is the Least Absolute Shrinkage and Selection Operator (Lasso) – see **Error! Reference source not found.** for details. A bootstrapping step with 1000 iterations was implemented to determine the uncertainty in the estimation of the mediation effects for each model. The mean, standard deviation, and confidence intervals values of the bootstrapped samples from the estimates were calculated and tested to identify the significant mediators. Indeed, the bootstrapping followed the design of the study, e.g., each treatment group was bootstrapped separately. The procedure was repeated for all the five different combinations of the modalities to complete our agnostic technique to explore the pathways. The results were further adjusted for multiple comparisons using the Bonferroni correction technique (see **Error! Reference source not found.** for further statistical information).

### Data Availability

Anonymised individual patient data is available on request, subject to data sharing agreement and UK research governance regulations.

## Results

In total, 90 patients gave consent and were randomised to each treatment arm, two of which did not complete a baseline MRI scan. After six months, complete fatigue follow-up, T1, resting state, and DTI scans were available for n=67 (see Table 1).

**Table 1:** Mean (SD) of the clinical characteristics of all the intervention/treatment group patients. The skewness values for the variables that have a mean/SD<2 are also reported.

|  | CBA<br>(N=21) | PEP<br>(N=24) | UC<br>(N=22) | Total<br>(N=67) |
| --- | --- | --- | --- | --- |
| **Gender** |  |  |  |  |
| Female | 14 (66.7%) | 19 (79.2%) | 18 (81.8%) | 51 (77.3%) |
| Male | 7 (33.3%) | 5 (20.8%) | 4 (18.2%) | 16 (22.8%) |
| **Age (years old)** | 62 (13) | 58 (13) | 58 (10) | 59 (12) |
| **Imaging site** |  |  |  |  |
| Aberdeen | 11 (52.4%) | 16 (66.7%) | 17 (77.3%) | 44 (65.7%) |
| Edinburgh | 10 (47.6%) | 6 (25%) | 5 (22.7%) | 21 (31.3%) |
| Glasgow | 0 | 2 (8.3%) | 0 | 2 (3%) |
|  | Mean (SD) = 11.30 (9.01) | Mean (SD) = 11.29 (10.55) | Mean (SD) = 12.06 (10) | Mean (SD) = 12.39 (10.39) |
|  | Median = 8.44 | Median = 8.86 | Median = 9.75 | Median = 9.37 |
| **Disease duration (years)** | IQR = 12.07 | IQR = 8.82 | IQR = 11.10 | IQR = 12.95 |
|  | Skewness = 0.86 | Skewness = 1.07 | Skewness = 1.40 | Skewness = 1.06 |
|  | Kurtosis = 2.71 | Kurtosis = 3.12 | Kurtosis = 4.62 | Kurtosis = 3.17 |
| **Baseline comorbidity index<sup>1</sup>** |  |  |  |  |
| 1 | 9 | 16 | 13 | 38 |

|  | **CBA**<br>(N=21) | **PEP**<br>(N=24) | **UC**<br>(N=22) | **Total**<br>(N=67) |
| --- | --- | --- | --- | --- |
| 2 | 6 | 7 | 8 | 21 |
| 3 | 3 | 1 | 1 | 5 |
| 4 | 2 | 0 | 0 | 2 |
| 7 | 1 | 0 | 0 | 1 |
| **RA disease activity<sup>2</sup> Baseline** | 4.5 (0.90) | 4.1 (1.1) | 3.6 (1.1) | 4.1 (1.1) |
| **RA disease activity 6 months** | 4.2 (0.97) | 4.2 (1.4) | 3.5 (1.2) | 4.0 (1.2) |
|  | Mean (SD) = 7.11 (7.06) | Mean (SD) = 7.84 (9.41) | Mean (SD) = 4.14 (2.46) | Mean (SD) = 6.22 (6.76) |
|  | Median = 4 | Median = 4 | Median = 4 | Median = 4 |
| **CRP (mg/l) Baseline** | IQR = 4 | IQR = 4.25 | IQR = 0 | IQR = 1.75 |
|  | Skewness = 2.97 | Skewness = 2.12 | Skewness = 1.65 | Skewness = 3.20 |
|  | Kurtosis = 12.70 | Kurtosis = 7.12 | Kurtosis = 6.66 | Kurtosis = 14.76 |
|  | Mean (SD) = 10.50 (13.02) | Mean (SD) = 6.16 (6.36) | Mean (SD) = 6.89 (16.36) | Mean (SD) = 6.80 (8.80) |
|  | Median = 4 | Median = 4 | Median = 4 | Median = 4 |
| **CRP (mg/l) 6 months** | IQR = 8 | IQR = 2 | IQR = 1.75 | IQR = 2 |
|  | Skewness = 2.28 | Skewness = 2.17 | Skewness = 4.76 | Skewness = 3.48 |
|  | Kurtosis = 7.61 | Kurtosis = 6.77 | Kurtosis = 24.13 | Kurtosis = 16.70 |
| **Chalder Fatigue Baseline** | 20 (6.8) | 21 (6.0) | 21 (4.6) | 21 (5.8) |
|  | Mean (SD) = 13.62 (7.17) |  |  |  |
| **Chalder Fatigue 6 months** | Median = 15 | 14 (7.3) | 19 (5.7) | 15 (6.8) |
|  | IQR = 7.50 |  |  |  |

| **CBA**<br>(N=21) | **PEP**<br>(N=24) | **UC**<br>(N=22) | **Total**<br>(N=67) |
| --- | --- | --- | --- |
| Skewness = -0.16 |  |  |  |
| Kurtosis = 2.51 |  |  |  |

<sup>1</sup>Charlson comorbidity index; <sup>2</sup>DAS 28; CBA = cognitive behavioural approach; CRP = C Reactive Protein; PEP = personalised exercise programme; RA = rheumatoid arthritis; UC = usual care

### Multi-modal MRI data curation

A combination of features was extracted from five different modalities of the original raw data: a) A total number of 84 volumetric features (values) were extracted from the MRI structural data. b) 68 area features (values), and c) 68 thickness features (values) were extracted from the MRI structural data. d) 3486 resting state functional connectivity features were extracted from the connectivity matrices of the rs-fMRI data. These matrices are diagonal, i.e., the relationship between one brain region and the other is the same regardless of the direction. e) 6972 structural connectivity features were extracted as from the connectivity matrices of the diffusion data.

### PEP mediation analysis

There were 17 structural and 13 functional connections identified as the mediators for fatigue changes for the PEP intervention group as compared to the usual care group, being the reference group (Figure 2 and appendix). There were no mediators identified for the volumetric metrics. The strongest structural connectivity mediators were left (L) isthmus cingulate cortex (ICC) to L paracentral lobule (PCL) with a mean mediation effect of −·46 (SD 0·61), L pars orbitalis to R paracentral with a mean mediation effect of −0·29 (SD 0·53), and the L lateral occipital gyrus to L cuneus with a mean mediation effect of −0·24 (SD 0·4). In terms of functional connectivity, the most significant mediators were the connections between L accumbens and R rostral anterior cingulate with a mean mediation effect of −0·62 (SD 0·61), the connection between L pallidum and R superior parietal lobule with a mean mediation effect of −0·41 (SD 0·5), and the connection between L pallidum and L inferior temporal gyrus with a mean mediation effect of −0·39 (SD 0·51).

**Figure 2:**
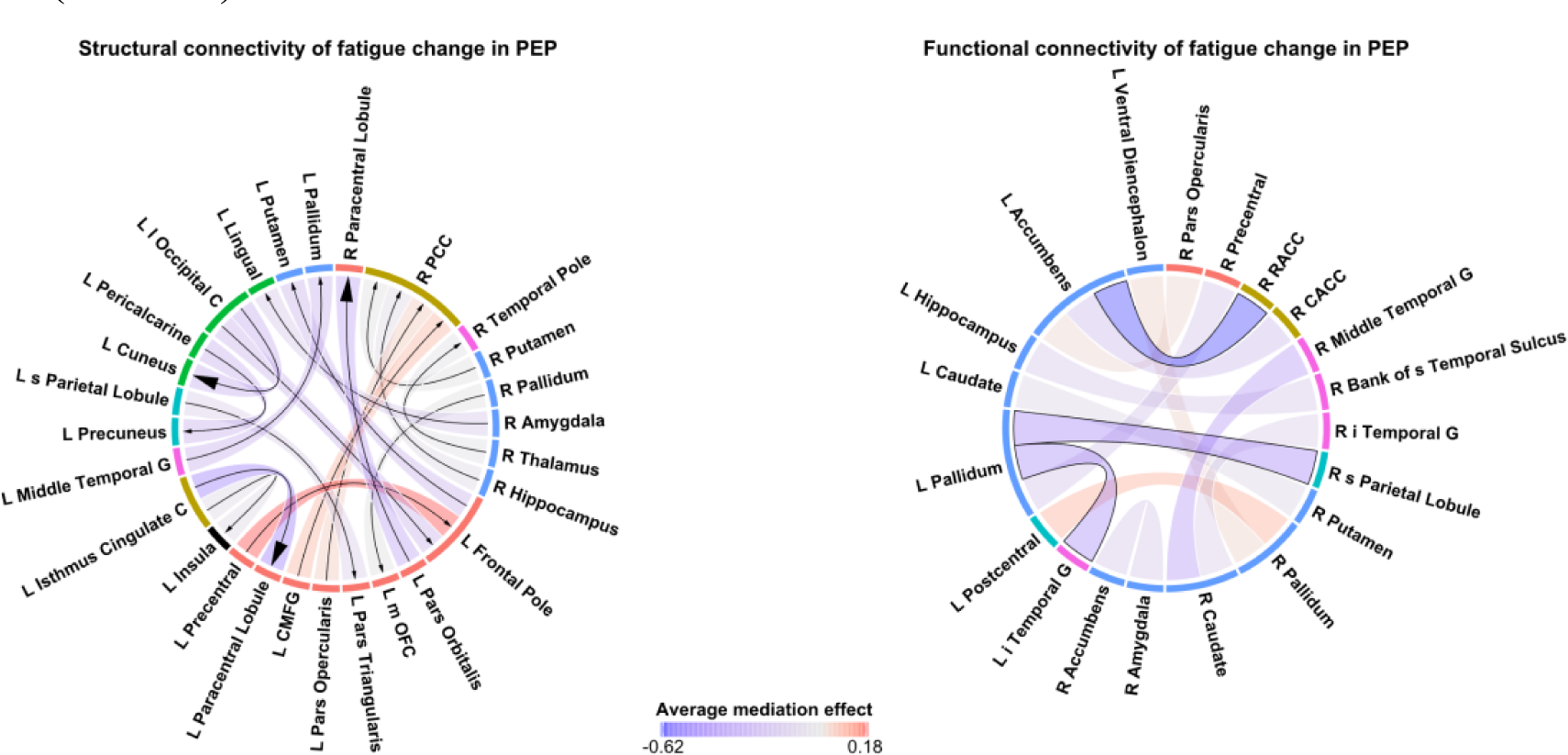
a) 17 structural connectivity features/connections (left) and b) 13 functional connectivity features/connections (right) were identified as the mediators for the fatigue changes in the PEP intervention group with the usual care group as the reference group in the analysis. The colour metric illustrates the average of the estimation of mediation effects from bootstrap samples (a total of 1000 iterations), which ranges from −0·62 to 0·18. The three strongest mediators are illustrated by bigger arrow heads and solid borders for structural connectivity and functional connectivity, respectively. The figure was created using ^25^. L/R = left/right. i/s = inferior/superior. m/l = medial/lateral. C = cortex. G = gyrus. L = lobule. OFC = orbitofrontal cortex. PCC = posterior cingulate cortex. RACC = rostral anterior cingulate cortex. CACC = caudal anterior cingulate cortex. CMFG = caudal middle frontal gyrus.

### CBA mediation analysis

17 structural and 12 functional connections were identified as mediators for CBA related fatigue change, relative to the usual care group (see Figure 3 and appendix). No mediators were identified for the volumetric modalities (volume, thickness, and area). The strongest mediators for the structural connections were L pars triangularis to L putamen with a mean mediation effect of −0·32 (SD 0·47), L-ICC to L-PCL with ß_Average_=-0·31 (SD 0·54), and the L accumbens to L transverse temporal gyrus with ß_Average_=-0·24 (SD 0·38). Although significant functional connections were identified, their size of effect was minimal.

**Figure 3:**
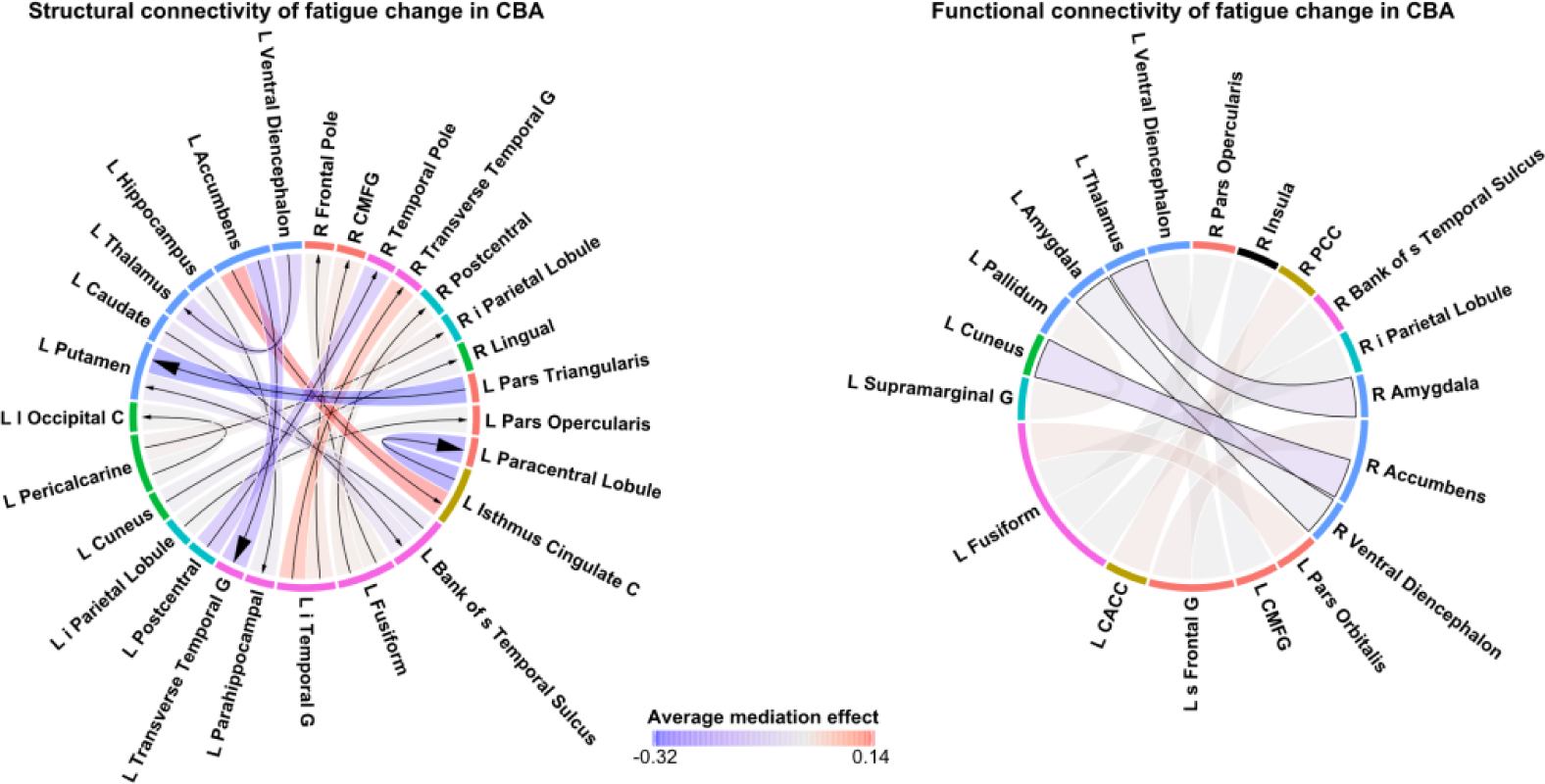
a) 17 structural connectivity features/connections (left) and b) 12 functional connectivity features/connections (right) were identified as the mediators for the fatigue changes in the CBA intervention group with the usual care group as the reference group in the analysis. The colour metric illustrates the average estimation of mediation effects from bootstrap samples (a total of 1000 iterations), which ranges from −0·32 to 0·14. The three strongest mediators are illustrated by larger arrow heads and solid borders for structural connectivity and functional connectivity, respectively. The figure was created using ^25^. L/R = left/right. i/s = inferior/superior. m/l = medial/lateral. C = cortex. G = gyrus. L = lobule. RACC = rostral anterior cingulate cortex. CACC = caudal anterior cingulate cortex. CMFG = caudal middle frontal gyrus.

The location of the most significant mediating connections, with an absolute effect size of above 0·1, for each intervention can be seen in Figure 4.

**Figure 4:**
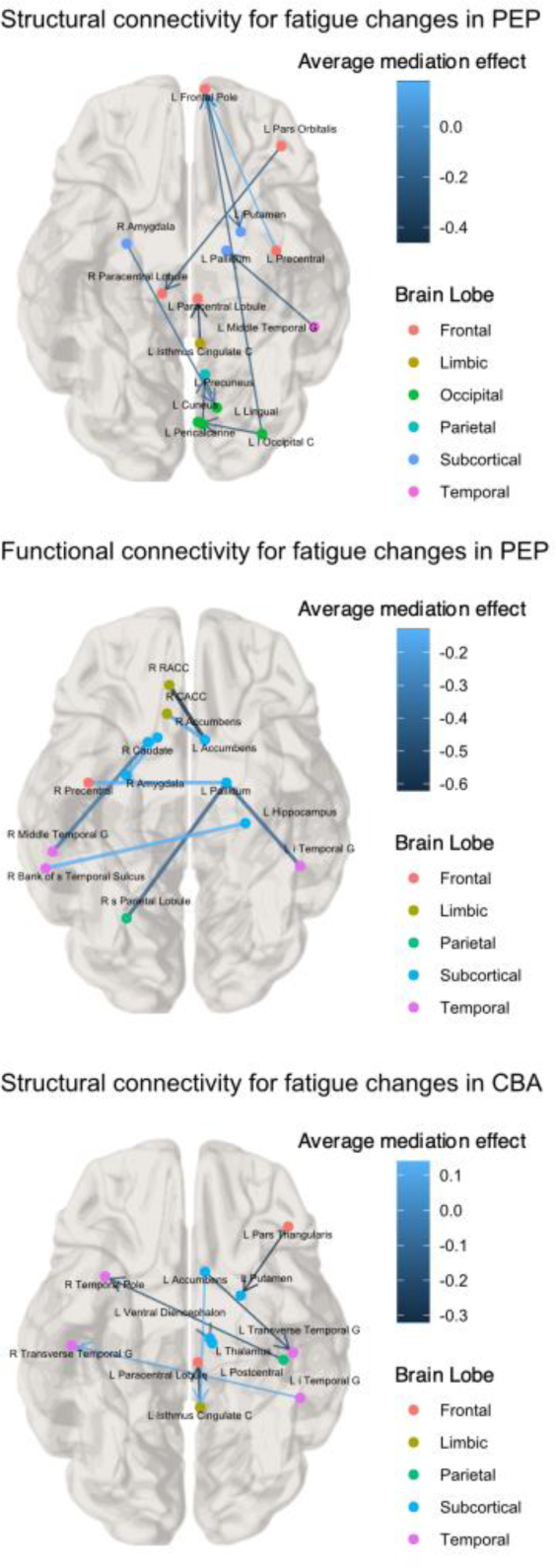
a) The location of the most significant structural connectivity features/connections (top) and b) The location of the most significant functional connectivity features/connections (middle) that were identified as mediators for fatigue change in the PEP intervention group. c) The location of the most significant structural connectivity features/connections (bottom) that were identified as mediators for fatigue changes in the CBA intervention group. The colour metric illustrates the average of the estimation of mediation effects from bootstrap samples (a total of 1000 iterations), which only includes the absolute values above 0·1. The figure was created using the brainconn software package ^26^. L/R = left/right. i/s = inferior/superior. m/l = medial/lateral. C = cortex. G = gyrus. L = lobule. RACC = rostral anterior cingulate cortex. CACC = caudal anterior cingulate cortex.

## Discussion

In this study, the first to investigate how exercise and psychosocial interventions alter the brain to improve RA-related fatigue, we have identified multiple white matter structural and functional brain connections that potentially mediated fatigue change. In contrast, individual brain structure volumetrics did not appear to have a causal role in symptom improvement.

Regarding structural connectivity, the effect of exercise on fatigue improvement was principally mediated by the white matter connection between the L-ICC and L-PCL. Notably, this specific feature was also a highly ranked mediator of the CBA intervention. This highlights the potential of this connection as a final common neurobiological substrate of fatigue, which both interventions appear to have successfully targeted. Although this is the first study to examine white matter connectivity in the context of RA fatigue mechanisms, we and others have previously employed DTI and similarly identified the ICC as a feature of fatigue in other inflammatory rheumatic diseases ^27,28^. Its role as a hub of the default mode network (DMN) may be highly relevant. DMN activity indicates introspective behaviour that may empower cognitive functions, but with overuse could lead to fatigue. Not only are such cognitions a common target of CBA, but in parallel, there is now extensive structural and functional MRI data evidencing a modulatory effect of exercise upon the DMN ^29^. The PCL is also a key hub of the somatosensory network (SMN), which was structurally related to fatigue in ankylosing spondylitis ^28^.

It is notable that both the DMN and SMN are critical networks in pain processing and a systematic literature review identified pain as one of the strongest predictors of RA fatigue ^30^. We have further showed that, unlike other common predictors, pain clustered with fatigue across almost all subjects.^31^. Thus, it is unsurprising that fatigue and pain appear to share neurobiological mechanisms, a hypothesis further supported by the known effectiveness of PEP and CBA in chronic pain conditions ^32^. Given the diverse natures of PEP and CBA, it may be perceived unusual that the ICC-PCL structural connection appears an important mechanism for both interventions. However, we know from the parent trial that many CBA recipients felt better able to increase their exercise levels once some of their cognitive challenges were addressed. There were no other neurobiological mediators shared between the interventions.

Overall, the neurobiological functional effect of PEP on fatigue, closely resembled our previous cross-sectional fMRI-based RA fatigue correlate findings ^9^, which strongly implicated overactivity of the DAN. In the current study, PEP appears to reduce the functional activity of connections involving the superior parietal lobule, middle temple gyrus and precentral regions (all landmarks of the DAN) with subsequent reductions in fatigue. Structural dysconnectivity of the precentral regions was also observed following PEP. More generally, across both interventions and MRI metrics, regions of the basal ganglia (pallidium, putamen, accumbens, caudate) were commonly identified, aligning with the canonical Chaudhuri & Behan model of chronic fatigue ^33,34^. This was originally framed on neurological observations of patients with lesions of the basal ganglia and their connections, especially Parkinson’s disease, where indeed fatigue is considered a primary manifestation ^35^. It is important to recognise that the identified candidate connections do not explain the totality of fatigue neural processing. The complexity of fatigue inevitably means that it will be underpinned by multiple regions/networks, however, our identified candidate connections represent critical components of the more expansive fatigue network and provides a focus for interventions.

One limitation of the present study was the inability to externally validate our findings since, to the best of our knowledge, there are no past or current randomised controlled clinical trials of RA with embedded multi-modal MRI brain imaging. However, our findings are biologically plausible, and have externally validated and prioritised neural correlates identified in previous studies ^8,9^. Further, our study did not identify volumetric features despite previous cross-sectional research highlighting their potential importance. This is potentially due to the longer timescale needed for volumetric changes to occur or be a consequence of differences in statistical power for these metrics not withstanding that this is the largest MRI brain study to date of any inflammatory rheumatic disease. This study also lacks alternative disease comparators and so although the highlighted brain regions have been consistently identified in other clinical populations, we are unable to confidently establish the transferability of these findings to other disease states. Finally, structural connectivity could be expected to provide an anatomical basis for function but in this study the overlap between the identified structural and functional connectivity mediators was small. However, a direct, spatially aligned, relationship is not always observed in other combined functional and structural connectivity MRI studies, rather in the context of pathology a more indirect relationship is proposed where the quality of the structural connectivity moderates functional activity ^36^.

Despite these shortcomings, this study discloses multiple novel routes for potential fatigue therapeutics. Neuromodulation techniques could non-invasively target identified regions using transcranial magnetic stimulation (TMS) or transcranial direct current stimulation (tDCS). These techniques affect regional brain functional and structural connectivity ^37,38^ through magnetic field inducing coils or scalp electrodes, respectively, and in severe depression their application to brain frontal regions is already established in routine clinical care ^39^. However, stimulation range limits TMS/tDCS targets to accessible regions on the surface of the brain and so precluding optimal modulation of our strongest candidate, the ICC-PCL white matter connection. Instead, the emerging application of transcranial pulse stimulation could overcome this limitation. Further, real-time neurofeedback paradigms could address functional targets by training patients to alleviate aberrant connectivity and potentially reduce subsequent fatigue ^40^.

## Conclusion

Understanding and managing fatigue presents one of the most sizeable contemporary challenges in the care of RA patients. Employing a gold standard causal analysis framework, this study examined the neurobiological mechanisms of action of two effective non-pharmacological RA fatigue interventions and in doing so identified and prioritised candidate brain substrates of RA fatigue which can now be targeted directly with a view to developing novel solutions for this unmet clinical need.

## Acknowledgements

We would like to thank all the investigators from the original LIFT study, including Kathryn Martin, Lorna Aucott, Neeraj Dhaun, Emma Dures, Stuart R. Gray, Elizabeth Kidd, Vinod Kumar, Karina Lovell, Graeme MacLennan, Paul McNamee, John Norrie, Lorna Paul, Jon Packham, Stefan Siebert, Alison Wearden, and Gary Macfarlane, without whose work this research would not be possible. Furthermore, we thank all the participants who generously supported the LIFT trial. We also acknowledge the contribution of the Trial Steering Committee and Data Monitoring Committee, and Brian Taylor and Mark Forrest (Centre for Healthcare Randomised Trials [CHaRT], University of Aberdeen, Aberdeen, UK) for their technical assistance.

## Patient and Public Involvement & Ethics Approval

All the details of the parent trial are published/included in BMJ Open paper doi: 10.1136/bmjopen-2018-026793.

## Data Sharing

Anonymised individual patient data will be made available following any reasonable request made to the corresponding author, subject to a data sharing agreement and UK research governance regulations. The intervention manuals can be found on https://www.abdn.ac.uk/iahs/research/epidemiology/lift-1286.php.

## Contributors’ Statement

**AD**: Methodology, Formal Analysis, Writing – Original Draft, Writing – Review & Editing; **SAW**: Methodology, Formal Analysis, Writing – Review & Editing; **KS**: Formal Analysis, Writing – Review & Editing; **SW, SR, JW, RE, EMB, & JC**: Writing – Review & Editing; **GW & NB**: Conceptualization, Methodology, Writing – Review & Editing, Supervision, Funding Acquisition. All authors approved the final version to be published.

## Key messages

Chronic Fatigue is a major clinical unmet need among patients with Rheumatoid Arthritis (RA). Current therapies are limited to non-pharmacological interventions, such as personalised exercise programmes (PEP) and cognitive behavioural approaches (CBA), however, still most patients continue to report severe fatigue.

Employing a gold standard causal analysis framework, this study examined the neurobiological mechanisms of action of two effective non-pharmacological RA fatigue interventions. To inform more effective therapies, we conducted an MRI brain study of PEP and CBA, nested within a randomised controlled trial (RCT), to identify their neurobiological mechanisms of fatigue reduction in RA.

The structural connection between the L-ICC and L-PCL appears to be a dominant mechanism for how both PEP/CBA reduces fatigue among RA patients. This supports its potential as a substrate of fatigue neurobiology and a putative candidate for future targeting.

## Footnotes

1 EULAR Disease Activity Score (DAS28), Charlson comorbidity index, peripheral measures of inflammation, Erythrocyte Sedimentation Rate and C Reactive Protein, pain, numerical rating scale assessing pain intensity, and fibromyalgia status, according to 2011 ACR survey criteria (baseline).

