## Appendix for "Characterising the neurobiological mechanisms of action of exercise and cognitive behavioural interventions for rheumatoid arthritis fatigue: an MRI brain study"

### The Mediation Analysis

The process of mediation analysis using this software package included three steps: (1) identifying potential mediators and transforming the data sets into the analytic format; (2) estimating the mediation effects based on the whole data; and (3) making inferences on mediation effects using a bootstrap method ^20^.

The method applied has numerous advantages. Most importantly, it allows an unlimited number of variables to be tested in the mediation analysis, either as mediators or covariates. Moreover, the variables can be of different types, e.g., continuous variables for mediators, multi-categorical variables for gender, etc. Furthermore, multiple mediators of different types are allowed in the pathway analysis simultaneously. Indirect effects transmitted by an individual mediator can be differentiated from the total effect, permitting the comparison of mediator importance. Finally, the mediation technique allows correlations among mediators.

#### Identifying Mediators

To be identified as a mediator, a variable must satisfy two conditions. First, the variable is significantly correlated with the predictor. To test this, we used chi-square test, ANOVA, or the Pearson’s correlation coefficient tests, depending on the variable types of the predictor and the potential mediator. The significance level (alpha2) is set to 0·1. This threshold was chosen to be lenient at the first stage and, also, to reduce the false negative rate going into the mediation effect size estimation, which is much more rigorous. The second condition is that the variable is significantly related with the outcome, given that all other related factors are included in the model. The significance level for this test is set to 0·1 (alpha). If both conditions are satisfied, the variable will be included in the data set as a mediator.

As previously mentioned, we used Least Absolute Shrinkage and Selection Operator (Lasso) as our regression technique since it is more suitable for the type of data that is analysed in this research. Lasso produces simpler and more interpretable models that incorporate only a reduced set of the predictors as compared to other methods, e.g., ridge. Furthermore, Lasso might perform better in a situation where some of the predictors have large coefficients, and the remaining predictors have very small coefficients (see ^41,42^ for further details). Additionally, Lasso reduces the number of variables, i.e., data reduction, and therefore tends towards not overfitting. Of note is that, in our analyses, the value of lambda was not varied arbitrarily, but was fixed for all the experiments, and therefore the models described in this research have fixed and known model parameters and hence, can be reproduced.

#### Statistical Inferences on Mediation Effects

We applied the bootstrap method to measure the uncertainty in estimating the mediation effects for each model. This involved calculating the variances and confidence intervals of the estimated mediation effects based on the estimated mediation effects from bootstrap samples. We implemented the bootstrapping with 1000 iterations. Then the average of the estimates is calculated from bootstrap samples (mean) along with the standard deviation (SD). A normal approximation technique was used to calculate the confidence intervals, with a significance level of 0·05 (alpha).

Later, we tested the confidence intervals to identify the significant mediators among the potential mediators. This was repeated for all the five different modalities to complete an agnostic approach to explore the pathways.

In brief, the relationship between the candidate mediating variables (brain imaging features) and the independent variable (intervention groups) and dependent variable (fatigue improvement, the outcome), after adjusting for the covariates, were tested for significance ^22,43,44^. Since we modelled each pathway with one candidate at a time, the brain imaging features were not included in any of the models as a covariate. This can be an option when a model with all the features is formed, in which case, if the candidate mediators only are related to the outcome and not the exposure variable, they will be included in the final model as covariates. To select the variables as potential mediators, a significant level of 0·1 for inclusion was applied ^22,43,44^. After the potential mediators were identified, a bootstrapping step was implemented to test the outcomes. 1000 iterations were applied, and the procedure was repeated for all the potential mediators individually. The results are further adjusted for multiple comparisons by applying the Bonferroni correction method.

One limitation of the *mmabig* package is that the generalised linear models were used to test the independence between variables and outcome. Therefore, linear relationships are assumed to identify mediators and covariates. The assumption can be relaxed by adapting the roughness of the concomitant rank test ^45^, a nonparametric method, to test the independence among variables. However, this is beyond the scope of this research.

#### Further Mediation Results

Table 2 to Table 5 depict the findings for the mediation analysis in details for all the different intervention combinations. In these tables, *mean* refers to the average of the estimation of mediation effects from bootstrap samples (a total of 1000 iterations). *sd* refers to the standard deviation of the estimation of mediation effects from bootstrap samples. *CI_upbd* and *CI_lwbd* refer to the upper bound and lower bound of the 95% confidence intervals built from the normal approximation method, respectively. The order of the connections in all the tables is by the absolute value (size) of the average of the estimation of mediation effects from bootstrap samples.

Table 2 - Mediation analysis results for structural connectivity mediators for the PEP intervention group with the usual care intervention group as the reference group.

| **From** | **to** | **mean** | **sd** | **95% CI** | |
| --- | --- | --- | --- | --- | --- |
|  |  |  |  | **L** | **U** |
| LH-Isthmus Cingulate Cortex | LH-Paracentral | -0·46 | 0·61 | -0·49 | -0·42 |
| LH-Pars Orbitalis | RH-Paracentral | -0·29 | 0·53 | -0·33 | -0·26 |
| LH-Lateral Occipital Gyrus | LH-Cuneus | -0·24 | 0·4 | -0·27 | -0·22 |
| LH-Middle Temporal Gyrus | LH-Pallidum | -0·23 | 0·42 | -0·26 | -0·21 |
| LH-Pericalcarine | LH-Precuneus | -0·23 | 0·41 | -0·25 | -0·2 |
| LH-Lateral Occipital Gyrus | LH-Frontal Pole | -0·21 | 0·36 | -0·23 | -0·18 |
| LH-Frontal Pole | LH-Putamen | -0·21 | 0·37 | -0·23 | -0·19 |
| LH-Precentral | LH-Frontal Pole | 0·18 | 0·34 | 0·16 | 0·2 |
| RH-Amygdala | LH-Lingual | -0·13 | 0·3 | -0·15 | -0·11 |
| LH-Superior Parietal Lobule | LH-Pars Triangularis | -0·098 | 0·24 | -0·11 | -0·083 |
| LH-Caudal Middle Frontal Gyrus | RH-Posterior Cingulate Cortex | 0·077 | 0·24 | 0·062 | 0·092 |
| LH-Pars opercularis | RH-Posterior Cingulate Cortex | 0·044 | 0·23 | 0·03 | 0·058 |
| RH-Hippocampus | RH-Posterior Cingulate Cortex | -0·043 | 0·21 | -0·056 | -0·03 |
| LH-Isthmus Cingulate Cortex | LH-Insula | -0·038 | 0·23 | -0·052 | -0·024 |
| RH-Thalamus Proper | RH-Temporal Pole | -0·021 | 0·22 | -0·034 | -0·0071 |
| RH-Putamen | RH-Posterior Cingulate Cortex | -0·015 | 0·19 | -0·027 | -0·0031 |
| RH-Pallidum | LH-Medial Orbito Frontal | -0·011 | 0·13 | -0·019 | -0·0029 |

Table 3 - Mediation analysis results for the functional connectivity mediators for the PEP intervention group with the usual care intervention group as the reference group.

| **from** | **to** | **mean** | **sd** | **95% CI** | |
| --- | --- | --- | --- | --- | --- |
|  |  |  |  | **L** | **U** |
| LH-Accumbens | RH-Rostral Anterior Cingulate | -0·62 | 0·61 | -0·65 | -0·58 |
| LH-Pallidum | RH-Superior Parietal Lobule | -0·41 | 0·5 | -0·44 | -0·38 |
| LH-Pallidum | LH-Inferior Temporal Gyrus | -0·39 | 0·51 | -0·42 | -0·36 |
| RH-Caudate | RH-Middle Temporal Gyrus | -0·33 | 0·49 | -0·36 | -0·3 |
| LH-Accumbens | RH-Caudal Anterior Cingulate Gyrus | -0·18 | 0·41 | -0·21 | -0·16 |
| LH-Pallidum | RH-Precentral | -0·14 | 0·31 | -0·16 | -0·12 |
| LH-Hippocampus | RH-Bank of the Superior Temporal Sulcus | -0·13 | 0·31 | -0·15 | -0·11 |
| RH-Amygdala | RH-Accumbens | -0·13 | 0·35 | -0·15 | -0·11 |
| RH-Caudate | RH-Inferior Temporal Gyrus | -0·099 | 0·31 | -0·12 | -0·079 |
| RH-Pallidum | LH-Postcentral | 0·086 | 0·22 | 0·072 | 0·1 |
| LH-Caudate | RH-Putamen | -0·057 | 0·24 | -0·072 | -0·042 |
| LH-Ventral Diencephalon | RH-Pallidum | 0·031 | 0·17 | 0·021 | 0·042 |
| LH-Accumbens | RH-Pars opercularis | 0·023 | 0·23 | 0·0084 | 0·037 |

Table 4 - Mediation analysis results for the structural connectivity mediators for the CBA intervention group with the usual care intervention group as the reference group.

| **from** | **to** | **mean** | **sd** | **95% CI** | |
| --- | --- | --- | --- | --- | --- |
|  |  |  |  | **L** | **U** |
| LH-Pars Triangularis | LH-Putamen | -0·32 | 0·47 | -0·35 | -0·29 |
| LH-Isthmus Cingulate Cortex | LH-Paracentral | -0·31 | 0·54 | -0·34 | -0·28 |
| LH-Accumbens | LH-Transverse Temporal Gyrus | -0·24 | 0·38 | -0·26 | -0·22 |
| LH-Postcentral | RH-Temporal Pole | -0·19 | 0·45 | -0·22 | -0·16 |
| LH-Ventral Diencephalon | LH-Thalamus Proper | -0·16 | 0·42 | -0·19 | -0·13 |
| LH-Accumbens | LH-Isthmus Cingulate Cortex | 0·14 | 0·32 | 0·12 | 0·16 |
| LH-Inferior Temporal Gyrus | RH-Transverse Temporal Gyrus | 0·1 | 0·31 | 0·085 | 0·12 |
| LH-Caudate | LH-Bank of the Superior Temporal Sulcus | -0·06 | 0·22 | -0·074 | -0·047 |
| LH-Bank of the Superior Temporal Sulcus | LH-Putamen | -0·056 | 0·2 | -0·068 | -0·044 |
| LH-Cuneus | RH-Lingual | -0·027 | 0·2 | -0·039 | -0·014 |
| LH-Inferior Temporal Gyrus | RH-Caudal Middle Frontal Gyrus | 0·026 | 0·21 | 0·013 | 0·039 |
| LH-Hippocampus | LH-Parahippocampal | -0·021 | 0·16 | -0·031 | -0·011 |
| LH-Fusiform | RH-Postcentral | 0·014 | 0·098 | 0·0084 | 0·021 |
| LH-Fusiform | RH-Frontal Pole | 0·01 | 0·075 | 0·0058 | 0·015 |
| LH-Pericalcarine | RH-Inferior Parietal Lobule | 0·0077 | 0·11 | 0·00099 | 0·014 |
| LH-Pericalcarine | LH-Lateral Occipital Gyrus | -0·0059 | 0·13 | -0·014 | 0·0019 |
| LH-Inferior Parietal Lobule | LH-Pars opercularis | 0·0017 | 0·15 | -0·0074 | 0·011 |

Table 5 - Mediation analysis results for the functional connectivity mediators for the CBA intervention group with the usual care intervention group as the reference group.

| **from** | **To** | **mean** | **sd** | **95% CI** | |
| --- | --- | --- | --- | --- | --- |
|  |  |  |  | **L** | **U** |
| RH-Accumbens | LH-Cuneus | -0·088 | 0·23 | -0·1 | -0·074 |
| LH-Thalamus Proper | RH-Amygdala | -0·053 | 0·45 | -0·081 | -0·025 |
| LH-Amygdala | RH-Ventral Diencephalon | -0·017 | 0·22 | -0·031 | -0·0034 |
| LH-Fusiform | LH-Pars Orbitalis | 0·017 | 0·12 | 0·0098 | 0·025 |
| LH-Caudal Anterior Cingulate Gyrus | RH-Posterior Cingulate Cortex | 0·012 | 0·081 | 0·0072 | 0·017 |
| LH-Caudal Middle Frontal Gyrus | RH-Insula | -0·0098 | 0·13 | -0·018 | -0·0016 |
| RH-Accumbens | LH-Superior Frontal Gyrus | 0·0089 | 0·073 | 0·0044 | 0·013 |
| LH-Pallidum | LH-Supramarginal Gyrus | 0·0068 | 0·11 | -0·00034 | 0·014 |
| LH-Fusiform | RH-Inferior Parietal Lobule | -0·0039 | 0·084 | -0·0091 | 0·0013 |
| LH-Fusiform | RH-Pars opercularis | 0·0012 | 0·067 | -0·0029 | 0·0054 |
| LH-Ventral Diencephalon | LH-Superior Frontal Gyrus | -0·0011 | 0·093 | -0·0068 | 0·0046 |
| LH-Fusiform | RH-Bank of the Superior Temporal Sulcus | 0·00079 | 0·16 | -0·0089 | 0·01 |
